# Mapping high-rate clusters of animal contact-related human *Salmonella enterica* single-state outbreaks in the United States, 2009–2022: A spatial epidemiological approach to inform public health surveillance

**DOI:** 10.64898/2026.04.04.26350168

**Authors:** Hammad Ur Rehman Bajwa, Suman Bhowmick, Csaba Varga

**Author notes:** **Corresponding author:** Dr. Csaba Varga, Varga Lab, Department of Pathobiology, College of Veterinary Medicine, University of Illinois Urbana-Champaign, Urbana, IL, USA. **E-mail address:** (C. Varga).

## Abstract

**Introduction:** Nontyphoidal *Salmonella enterica* (NTS) is a major zoonotic enteric pathogen. Animal contact-related NTS outbreaks have increased in the United States of America (U.S.) over the last decade. Geospatial analysis can identify locations with elevated risk of NTS outbreaks where public health authorities can focus their NTS prevention and intervention efforts.

**Methods:** We analyzed NTS outbreak data reported from individual states to the Centers for Disease Control via the National Outbreak Reporting System between 2009 and 2022 across the continental contiguous U.S. A geospatial analytical framework that included disease mapping, spatial interpolation, and global and local clustering methods was applied to identify regions with high NTS outbreak rates.

**Results:** A total of 104 NTS single-state outbreaks were reported to the National Outbreak Reporting System (NORS) during the study period. The mean annual incidence rate was 0.02 NTS outbreaks per million person-years. The primary animal contact categories associated with these outbreaks were mammals (cattle, pigs, sheep, and horses), birds (backyard chickens, ducklings, and turkeys), and reptiles (turtles and lizards). Exposure settings included farms, fairgrounds, agricultural feed stores, veterinary clinics, dairy/agricultural settings, and residential settings. The local cluster detection methods consistently identified areas with significantly high NTS animal contact-related outbreak rates in the Mountain West, Midwest, and Northeast of the US.

**Conclusion:** NTS animal contact-related single-state outbreaks revealed distinct spatial clustering across the United States, with potentially higher risks in the Mountain West, Midwest, and Northeast. Diversity of animal-contact sources and exposure settings depicted complex transmission dynamics of NTS. Focused prevention and control programs in these areas are needed to mitigate the burden of NTS outbreaks.

**Impacts:**

- The most common animals linked to outbreaks were mammals, birds, and reptiles
- Main exposure settings included farms, veterinary clinics, feed stores, shelters, schools, and homes.
- Outbreaks clustered in the Mountain West, Midwest, and Northeast regions.
- Region-specific prevention and interventions are needed to reduce the burden of illnesses.

## 1. Introduction

Nontyphoidal Salmonella enterica (NTS) are an important bacterial enteric pathogen that impacts human health (Majowicz et al., 2010). Globally, NTS are a leading cause of gastroenteric infections, with an estimated 153.1 million cases and 56,969 deaths (Kirk et al., 2015). NTS is primarily transmitted through the consumption of contaminated food products; however, animal contact-related NTS outbreaks are also significant. Animal contact-associated salmonellosis occurs through direct contact with infected animals or indirect contact with their contaminated environment (Hoelzer et al., 2011).

Since NTS are zoonotic pathogens with diverse animal reservoirs, they may exhibit regional variation in transmission patterns and disease burden (Yates et al., 2025). Regional differences may exist in agricultural practices, livestock densities, animal-contact exposure types, urbanization, environmental factors, and demographic and socioeconomic characteristics of human populations, which can influence NTS transmission (Varga et al., 2013; John et al., 2025; Yates et al., 2025). Understanding how these factors impact NTS transmission is important to designing local public health prevention and intervention programs.

Spatial analysis is useful in understanding the NTS distribution across space and time, identifying high-risk clusters, and assessing risk factors (Seixas et al., 2018). Several studies have been conducted to understand the distribution and clustering of NTS. In Italy, NTS cases clustered in the southern part of the country, where the livestock population was lower, suggesting a non-livestock environmental NTS reservoir (Graziani et al., 2015). Similarly, in the Netherlands, NTS incidence was not spatially associated with exposure to livestock, suggesting that foodborne transmission was the main exposure source (Mulder et al., 2024). A study in Portugal found persistent urban clusters, highlighting the importance of demographic factors in shaping NTS transmission (Seixas et al., 2018). In Canada, studies have examined the impact of socioeconomic (Varga et al., 2013a; John et al., 2025) and demographic factors (Varga et al., 2013b), and living close to a slaughterhouse (Paphitis et al., 2021) on NTS incidence. Other Canadian studies assessed spatial-temporal clustering of NTS infections, identifying high-rate clusters during warmer months and highlighting regional variability in infection rates (Varga et al., 2015; Valcour et al., 2016; John et al., 2024).

In the United States of America (US), the Centers for Disease Control and Prevention (CDC) coordinates national enteric disease surveillance through the National Outbreak Reporting System (NORS), which collects foodborne, waterborne, person-to-person transmitted, environmental contamination, and animal contact outbreaks reported by state, local, and territorial public health agencies (CDC, 2023). Previous research using NORS data has compared the characteristics of foodborne and animal-contact-related NTS outbreaks, identifying that children are more affected in animal-contact-related outbreaks. These NTS outbreaks were often linked to contact with infected backyard poultry, petting zoos, or reptiles (Marus et al., 2019). Other national-level analyses found that NTS was the second most reported enteric pathogen and had the second highest hospitalization rates among outbreak-associated infections (Wikswo et al., 2022). In addition, a longitudinal national study of Salmonella Enteritidis outbreaks from 1990 to 2015 revealed regional differences in their incidence and exposure sources (Sher et al., 2021). A Florida-based study identified persistent high NTS infection incidence areas in the northern part of the state (Li et al., 2021).

Despite the extensive literature on NTS infections, recent studies from the US have primarily focused on specific NTS serovars, short study periods, or narrowly defined geographic regions. A gap remains in national-level analyses examining longitudinal spatial and spatio-temporal clustering of animal-contact-associated NTS outbreaks.

To address this knowledge gap, the present study uses the data from the NORS to apply a geospatial analytical framework and assess the distribution and clustering of animal-contact-related single-state and multi-state NTS outbreaks reported at the state level in the United States (US) from 2009 to 2022. Findings from this study can inform regionally focused public health strategies aimed at reducing zoonotic transmission of NTS and provide a model for analyzing other enteric pathogens.

## 2. Materials and Methods

### 2.1. Study setting

All animal contact-related NTS outbreak information that was reported to the CDC via NORS between 2009 and 2022 was obtained through a data request. Single-state outbreaks, where exposures occurred in a single state, were included, and because of the geospatial analyses, we included single-state outbreak data from the contiguous U.S., which consists of 48 states and the district of Columbia that share a land border, and excludes Alaska, Hawaii, and U.S. territories and possessions (American Samoa, Guam, Puerto Rico, and the U.S. Virgin Islands) (**Figure S1)**. An outbreak was defined as the occurrence of more cases than expected within a period, a specific location, and a target population (Reintjes and Zanuzdana, 2009). State-level population data for the study period were obtained from the U.S. Census Bureau (U.S. Census Bureau, 2024a). The administrative boundary shapefile was obtained from the TIGER/Line database (U.S. Census Bureau, 2024b). For all spatial analysis, the shape file was projected to the USA Contiguous Albers Equal Area Conic USGS, as it is well-suited for areas extending in an east-to-west orientation (Esri, 2025).

### 2.2. Data analysis

The data cleaning, management, and descriptive statistics were performed using the R statistical software (Version 4.1.2 (2021-11-01)) (R Core Team, 2021). The spatial analyses were conducted in ArcGIS Pro 10.7.1 (Environmental Systems Research Institute, Inc., Redlands, CA, USA).

#### 2.2.1. Mapping nontyphoidal *Salmonella* single-state outbreak incidence rates

In the first step, state-level NTS single-state outbreak mean incidence rates (IRs) per 1 million population-years (1 MPY) were calculated for two periods (2009 – 2015 and 2016 – 2022) by dividing the number of NTS outbreaks in each state in each period by the corresponding state and period-specific population and multiplying it by 1 million. The state-level mean IRs per 1 MPY for the two periods were illustrated in choropleth maps, using the natural breaks classification with 5 breaks (Jenks et al., 1967). The categorization was slightly changed to include a class representing an IR of 0.

In the next step, to account for unstable IRs of states with small populations, the spatial empirical Bayes (SEB) smoothing method was used (Marshall, 1991) in GeoDa software (Anselin et al., 2006). For spatial weighting, the 1^st^-order Queen contiguity criterion was used, which defines the neighbors as spatial units with a shared vertex or edge, and considers the immediate neighbors (Anselin et al., 1996). The distribution of raw (non-smoothed) and SEB-smoothed NTS outbreak IRs was illustrated in choropleth maps.

In the last step, empirical Bayes kriging (EBK) (Gribov and Krivoruchko, 2020), was used to spatially interpolate the NTS IRs. The EBK method estimates IRs empirically from the existing data and averages IR predictions over multiple simulations to capture uncertainty in the data (Krivoruchko et al., 2019). The results of the spatial interpolation of the NTS IRs for the two study periods were illustrated in isopleth maps.

#### 2.2.2. Global cluster analysis

For two time periods (2009 – 2015 and 2016 – 2022), the incremental spatial autocorrelation analysis (ISA) using Moran’s I method estimated global clustering of NTS IRs over a series of increasing incremental Euclidean distances (Mitchell, 1999). For each distance increment, z-scores and p-values were calculated and illustrated in a graph, and the highest z-score represented the maximum peak, where spatial processes promoting spatial clustering were more active (Mitchell and Scott Griffin, 2021). The distance band associated with the highest z-score was selected for the subsequent local cluster analyses.

#### 2.2.3. Local cluster analysis

Each state was represented by a polygon, its centroid, and the corresponding IR for the respective study periods (2009 – 2015 and 2016 – 2022). For the conceptualization of spatial relationships, the “zone of indifference” parameter, which assigned equal spatial weights to states within the specified threshold distance around the index area, while this weight decreased as that distance was passed, allowing for a gradual decay of spatial influence (Mitchell, 1999).

##### 2.2.3.1. Hot spot analysis (Getis-Ord Gi* statistics)

The hotspot analysis identified statistically significant (p≤0.05) clusters of high or low NTS outbreak IRs. The statistically significant high z-score depicted a hot spot, where a high IR state was surrounded by high IR states, a low z-score depicted a cold spot, where a state with low IR was surrounded by low IR states, while an almost zero z-score revealed no significant clustering and implied random distribution (Getis and Ord, 1992).

##### 2.2.3.2. Cluster and outlier analysis (Anselin’s local Moran’s I)

This method detects clusters with similar IRs and spatial outliers where IRs differ significantly from the neighboring states. Four cluster types are identified: High-High (high IR state surrounded by High IR states), Low-Low (low IR state surrounded by low IR states), High-Low (a high IR state surrounded by low IR states), and Low-High outlier (low IR state surrounded by high IR states) (Anselin, 1995).

#### 2.2.4. Scan statistics

Scan statistics were conducted using the SaTScan v10.3.2 software (Kulldorff et al., 2022) to identify purely spatial and space-time clusters of NTS single-state outbreaks across the US during the whole study period from 2009 to 2022. The retrospective discrete Poisson model was applied (Kulldorff et al, 1997). For the spatial scan analysis, the circular scanning window was set to include up to 10% of the population at risk (Kulldorff et al., 2022), which searched for high or low NTS IR clusters. The retrospective space-time analysis included a cylindrical scanning window (a geographic base with a temporal interval height) that searched for high or low rate NTS IR clusters. The maximum spatial cluster size was set to include 10 percent of the population at risk, and 50 percent of the study period. The relative risk (RR) for each identified cluster was estimated as the ratio of observed to expected outbreaks inside the cluster relative to outside (Kulldorff et al., 2022). The RR was calculated as,

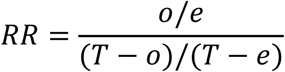

Where *o* was the total number of NTS single-state outbreaks in a state, *e* was the total number of expected NTS single-state outbreaks in a state, *T* was the total number of observed NTS single-state outbreaks in the US.

An RR greater than 1 indicated a higher-than-expected while RR<1 reflected a lower-than-expected NTS outbreak (Kulldorff et al, 2022). To avoid the assumption of identical risk within a significant cluster, RR for each area included in the cluster was calculated and illustrated in maps. The statistical significance was determined using Monte Carlo hypothesis testing with 999 replications (Kulldorff et al, 2022).

## 3. Results

### 3.1. Mapping raw NTS outbreak incidence rates

Figure 1 (A and B) illustrates the state-level raw IRs of NTS outbreaks for the two study periods (2009 – 2015 and 2016 – 2022) in choropleth maps.

**Figure 1.**
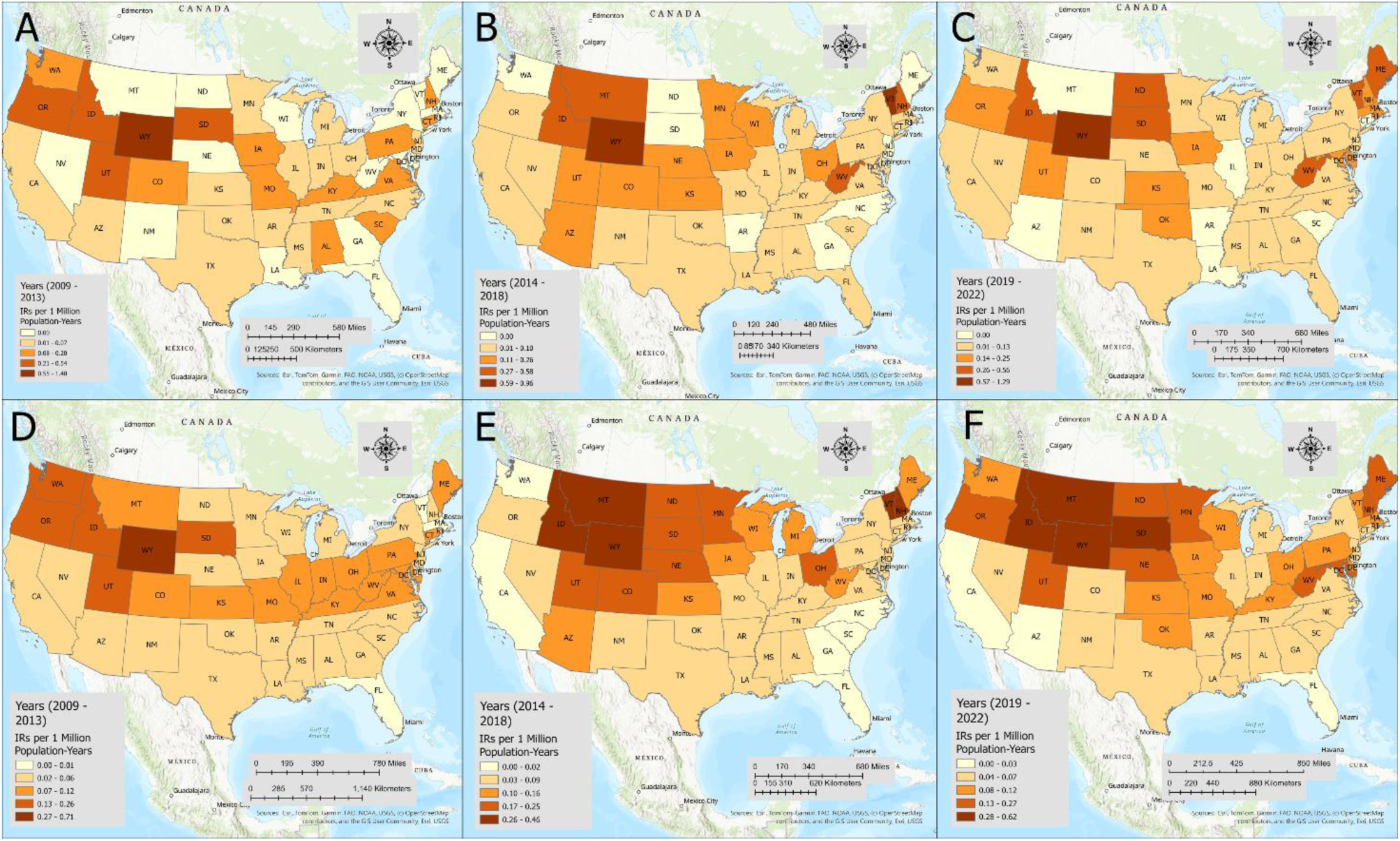
Incidence rates (per 1 million population-years) of single-state outbreaks across the U.S. for two periods (2009 – 2015, and 2016 – 2022) (Raw incidence rates: A and B; Smoothed incidence rates: C and D). The spatial empirical Bayes smoothing with first-order queen contiguity weights was used to smooth the incidence rates.

In the first period, Wyoming reported the highest IR (1.00 per 1MPY), followed by Vermont (0.46), New Hampshire (0.21), Alaska (0.20), Utah (0.20), South Dakota (0.18), and Colorado (0.16). The states with moderate to low IRs were Arkansas (0.05), Minnesota (0.05), Michigan (0.03), Arizona (0.02), and Illinois and North Carolina (0.01). The states that had no reported outbreaks during this period were Alabama, Hawaii, Idaho, Kansas, Montana, New Mexico, Tennessee, Texas, and West Virginia (Table S4 and Figure 1).

During the second period, Idaho reported the highest IR (0.32 per 1MPY), followed by Wyoming (0.25), West Virginia (0.24), Utah (0.18), Nebraska (0.15), Montana (0.14), New Hampshire (0.11), Hawaii (0.10), and Wisconsin (0.05). The states with moderate to low IRs were Michigan (0.04), Colorado (0.03), Maryland (0.02), and New York (0.01). The states reported no outbreaks during this period were Alaska, Kentucky, Oregon, Arkansas, Illinois, Missouri, and Virginia (Supplemental Table S3 and Figure 1).

### 3.2. Mapping smoothed NTS outbreak incidence rates

The NTS outbreaks’ SEB smoothed IRs in each state were presented in choropleth maps for the two study periods (2009 – 2015 and 2016 – 2022) in Table S2 and Figure 1. In the first period (2009 – 2013), Wyoming reported the highest IR of 0.070 per million population-years, followed by Vermont (0.03) and Utah (0.02). The states with the lowest IRs were Nebraska (0.01), New Mexico (0.006), New York (0.002), and Texas (0.0008). In the period 2016 – 2022, Utah reported the highest IR of 0.02, followed by Wyoming (0.01), Minnesota (0.009), and Ohio (0.006). The states with the lowest IRs were Texas (0.0008), Oregon (0.0007), and Virginia and Washington (0.0004) (Table S4 and Figure 1).

### 3.3. Empirical Bayes kriging of NTS outbreak incidence rates

Figure 2 illustrates the spatially interpolated IR values in isopleth maps for the two study periods. During 2009 – 2015, the elevated interpolated values (0.020 – 0.046) were observed in the Mountain West region. Moderate to low values were observed in the upper Northeast and upper Midwest region (0.0045 – 0.020).

**Figure 2.**
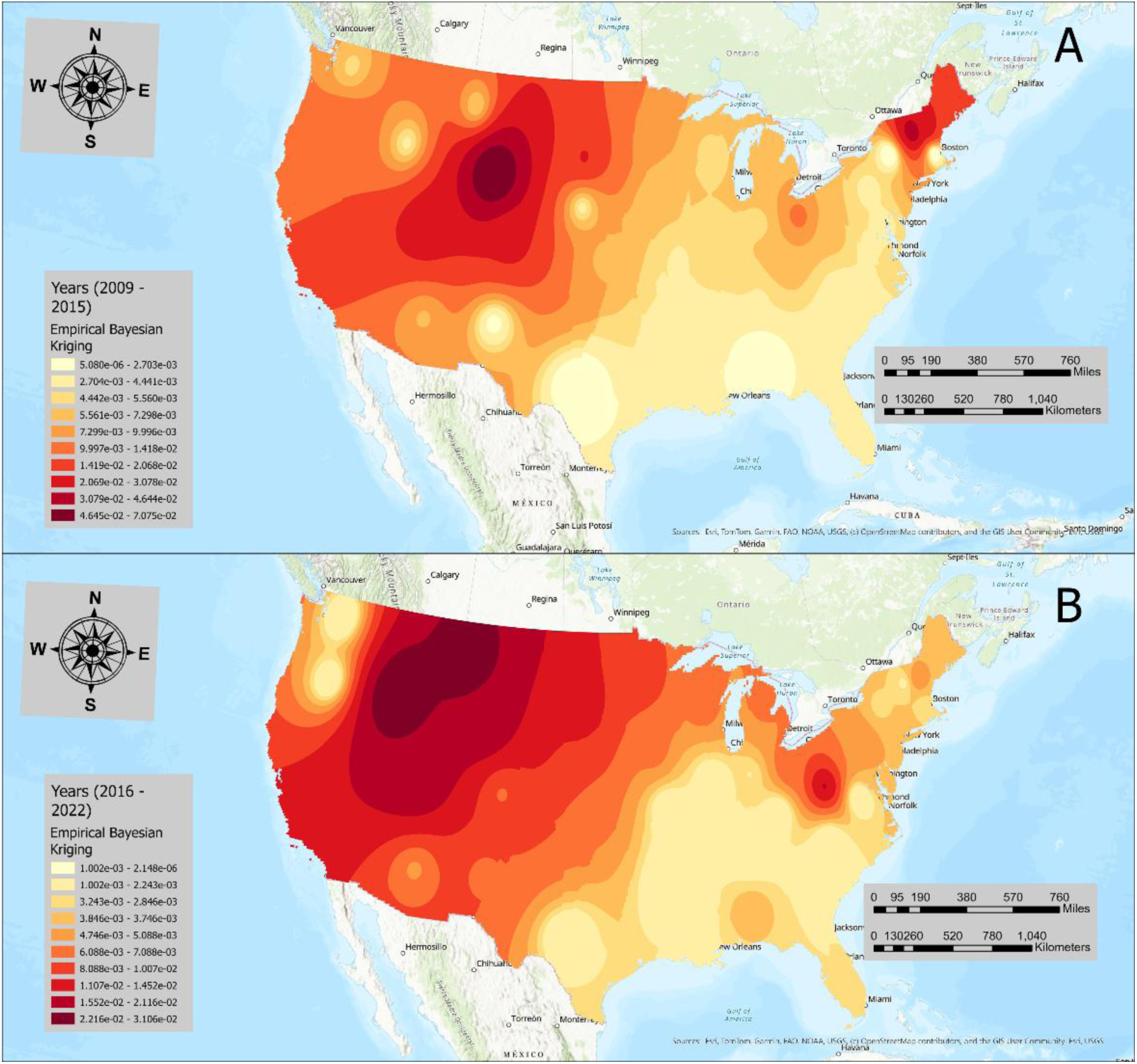
Spatial interpolation of outbreak incidence rates for two periods (2009 – 2015, and 2016 – 2022) across the U.S. The empirical Bayesian kriging method was used to interpolate the outbreak incidence rates (outbreaks per 1 million person-years).

The lowest interpolated values were in the Southern and Southwestern regions. During the second period (2016–2022), the Mountain West and parts of the upper Northeast emerged as high-interpolated value regions (0.011–0.0031), and this trend further radiated towards the Pacific midwestern region. Moderate to low values were observed in the Central Plains and the lower Northeastern region (0.005 – 0.011), while various parts of the Southern region were consistently lowest (Figure 2).

### 3.4. Global clustering

The incremental autocorrelation analysis assessed the global clustering of NTS IRs across the US for two study periods: 2009-2015 and 2016-2022 (Figure 3).

**Figure 3.**
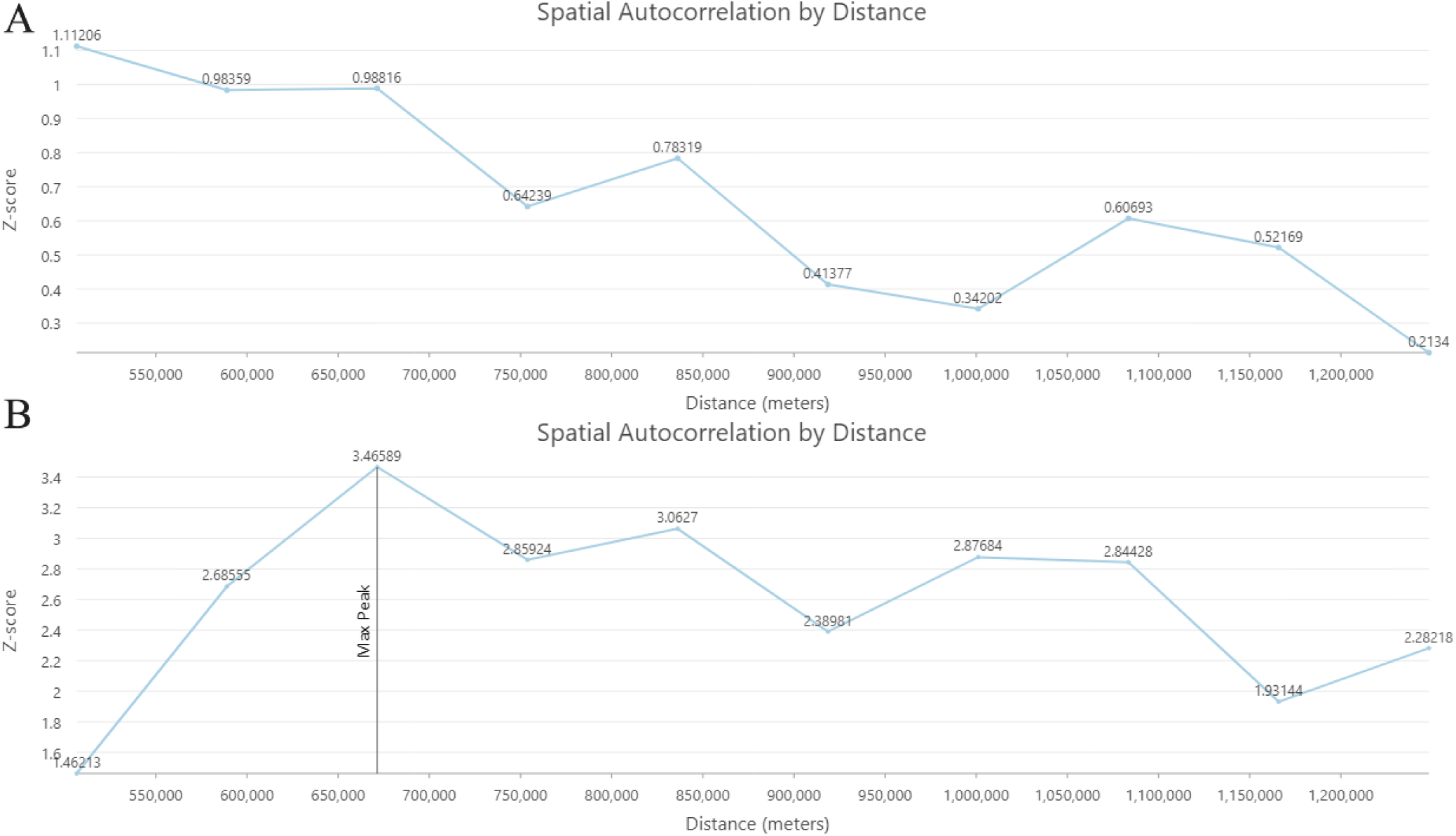
Incremental spatial autocorrelation analysis of outbreak incidence rates for two periods (2009 – 2015, and 2016 – 2022) across the U.S. Ten distance increments were used with the fixed-distance band to conceptualize spatial relationships. Significant at p≤0.05, Z-score ≥1.96.

The analysis estimated significant spatial autocorrelation across 10 incremental distances using global Moran’s I statistic. In 2009 – 2015, no significant peak was observed (Figure 3 and Table S5). While in the period 2016 – 2022, the first and maximum peak was observed at 671.38 km (Moran’s I = 0.252, z = 3.46, p-value = 0.001) (Figure 3 and Table S6).

### 3.5. Local clustering

#### 3.5.1. Hot spot (Getis-Ord Gi*) analysis

In 2009 – 2015, significant hotspots were detected in Wyoming, Montana, and Colorado (99% confidence) (Fig 4, A and B).

**Figure 4.**
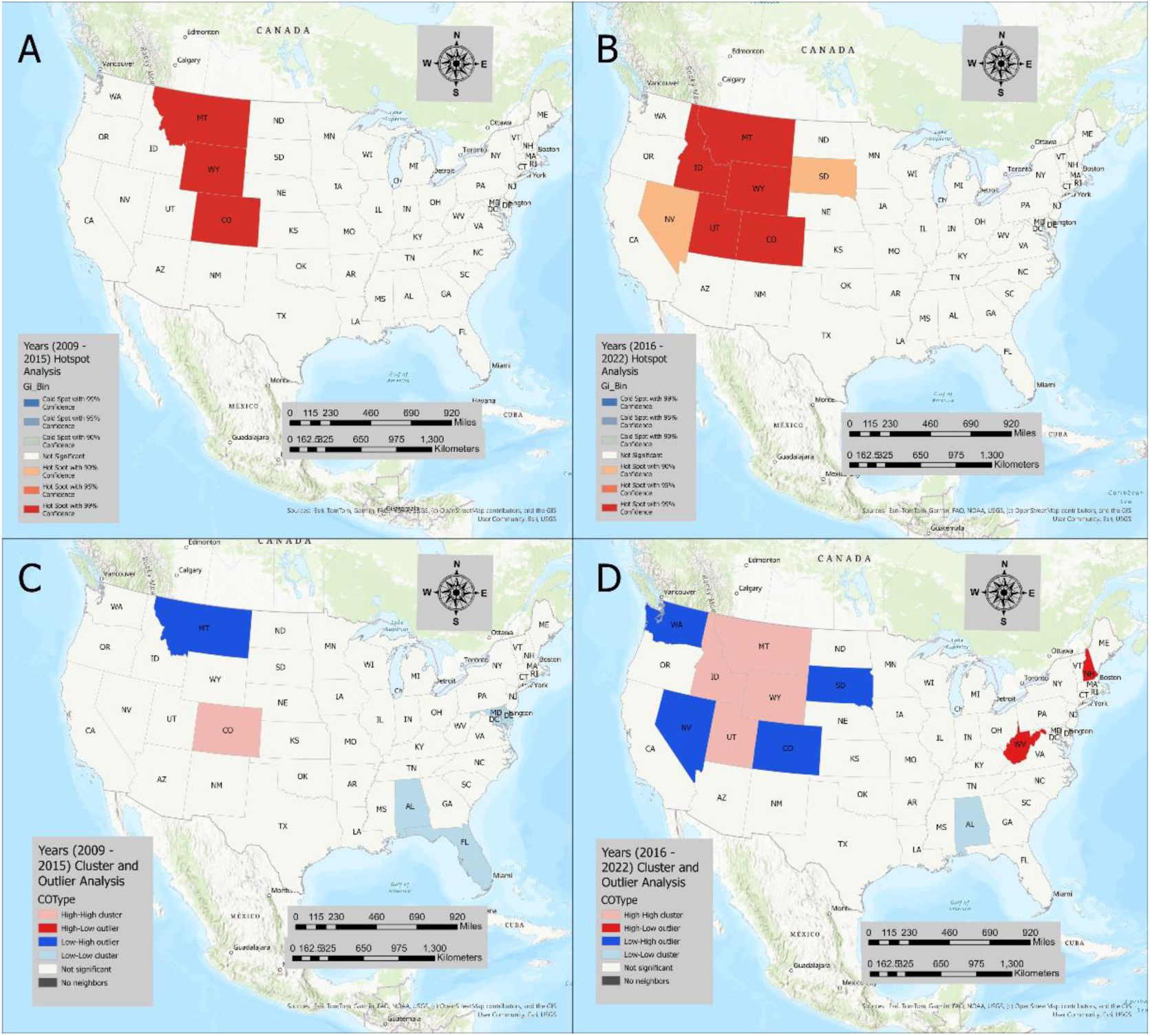
Maps illustrating the results of the local cluster analysis of outbreak incidence rates for two periods (2009 – 2015, and 2016 – 2022) across the U.S. Hotspot (Getis-Ord Gi*) analysis (A and B) and Cluster, and Outlier analysis (Anselin Local Moran’s I) (C and D). The animal contact sources that were associated with NTS single-state outbreaks in these states during this period were baby chicks, ducklings, small turtles, pigs, and small mammalian pets. The most common exposure settings were “Others” (Table S1 in the Supplement). During 2016 – 2022, Wyoming, Montana, Idaho, Colorado, and Utah were detected as hotspots (99% confidence), followed by Nevada and South Dakota as hotspots with (90% confidence). While no cold spots were detected during this period (Figure 4). The animal contact sources that were associated with these states during this period were poultry (chicks, ducklings, turkeys, and chickens) and mammals (cattle, pigs, horses, and mice), while the exposure settings were “Others”, fairground, farm/dairy/agricultural settings, and veterinary clinic (Table S1).

#### 3.5.2. Cluster and outlier analysis (Anselin Local Moran’s I)

Figure 4 (C and D) illustrates the significant clusters identified by the local Moran’s I statistic. In the first period (2009 – 2015), significantly high-high (HH) clusters were identified only in Colorado, and low-low (LL) clusters were detected in Alabama and Florida. The LH outliers were detected only in Montana; there were no HL outliers during this period (Figure 4).

The associated animal contact sources linked to NTS outbreaks in these states during this period were baby chicks, ducklings, small turtles, and small mammalian pets. The exposure settings were “Others” (Table S1).

During the second period (2016 – 2022), HH clusters were found in Montana, Idaho, Utah, and Wyoming, and an LL cluster in Alabama. The HL outliers were detected in West Virginia and New Hampshire. LH outliers were found in Washington, Nevada, Colorado, and South Dakota during this period (Table S1 and Figure 4). The animal contact NTS outbreak exposure sources that were associated with these states during this period were poultry (chicks, ducklings, turkeys, and chickens), mammals (cattle, pigs, sheep, goat, horse and ferret), and reptiles (turtles and lizards), while the exposure settings were “Others”, farm/dairy/agricultural settings, agricultural feed store, and Child daycare/preschool (Table S1).

### 3.6. Scan Statistics

In purely spatial analysis, three significant high-rate spatial clusters were detected using a discrete Poisson model (Table 1 and Figure 5).

**Table 1.** Spatial and space-time clusters of animal contact-related NTS outbreaks across the U.S., 2009 – 2022.

| Pathogen | Cluster | Model type | Number of States | Time frame | Population | N/C | E/C | O/E | LR | RR | p-value |
| --- | --- | --- | --- | --- | --- | --- | --- | --- | --- | --- | --- |
| Nontyphoidal *Salmonella* (NTS) | 1. | Purely spatial | 7 | – | 14580728 | 28 | 6.62 | 4.23 | 21.70 | 5.51 | 0.001 |
|  | 2. | Purely spatial | 2 | – | 13464767 | 16 | 6.11 | 2.62 | 6.05 | 2.93 | 0.04 |
|  | 3. | Purely spatial | 1 | – | 19616550 | 1 | 8.90 | 0.11 | 6.05 | 0.10 | 0.04 |
|  | 4. | Purely spatial | 2 | – | 1975331 | 5 | 0.90 | 5.58 | 4.57 | 5.82 | 0.176 |
|  | 5. | Purely spatial | 2 | – | 19433159 | 2 | 8.82 | 0.23 | 4.10 | 0.21 | 0.266 |
|  | 6. | Purely spatial | 2 | – | 15020643 | 5 | 6.82 | 0.29 | 2.48 | 0.28 | 0.830 |
|  | 7. | Purely spatial | 4 | – | 20597223 | 5 | 9.35 | 0.53 | 1.32 | 0.51 | 0.997 |
|  | 1. | Space-time | 4 | 2012<br>–<br>2018 | 10782518 | 17 | 2.42 | 7.01 | 19.69 | 8.26 | 0.001 |
|  | 2. | Space-time | 2 | 2013<br>–<br>2018 | 13464767 | 13 | 2.62 | 4.97 | 11.03 | 5.57 | 0.005 |
|  | 3. | Space-time | 3 | 2014<br>–<br>2019 | 17170774 | 14 | 3.35 | 4.18 | 9.97 | 4.70 | 0.01 |
|  | 4. | Space-time | 2 | 2013<br>–<br>2017 | 1975331 | 5 | 0.32 | 15.72 | 9.20 | 16.50 | 0.02 |
|  | 5. | Space-time | 1 | 2009<br>–<br>2009 | 577146 | 2 | 0.018 | 109.52 | 7.43 | 111.76 | 0.09 |
|  | 6. | Space-time | 2 | 2016<br>–<br>2022 | 19451717 | 0 | 4.54 | 0 | 4.65 | 0 | 0.77 |
|  | 7. | Space-time | 1 | 2009<br>–<br>2015 | 19616550 | 0 | 4.43 | 0 | 4.53 | 0 | 0.82 |
|  | 8. | Space-time | 2 | 2015<br>–<br>2021 | 19433159 | 0 | 4.42 | 0 | 4.51 | 0 | 0.83 |
|  | 9. | Space-time | 3 | 2009<br>—<br>2014 | 16459085 | 0 | 3.10 | 0 | 3.15 | 0 | 0.999 |

<sup>1</sup>N/C – Number of cases, E/C – Expected cases, O/E – Observed/Expected, LR – Likelihood
ratio, and RR – Relative Risk.

**Figure 5.**
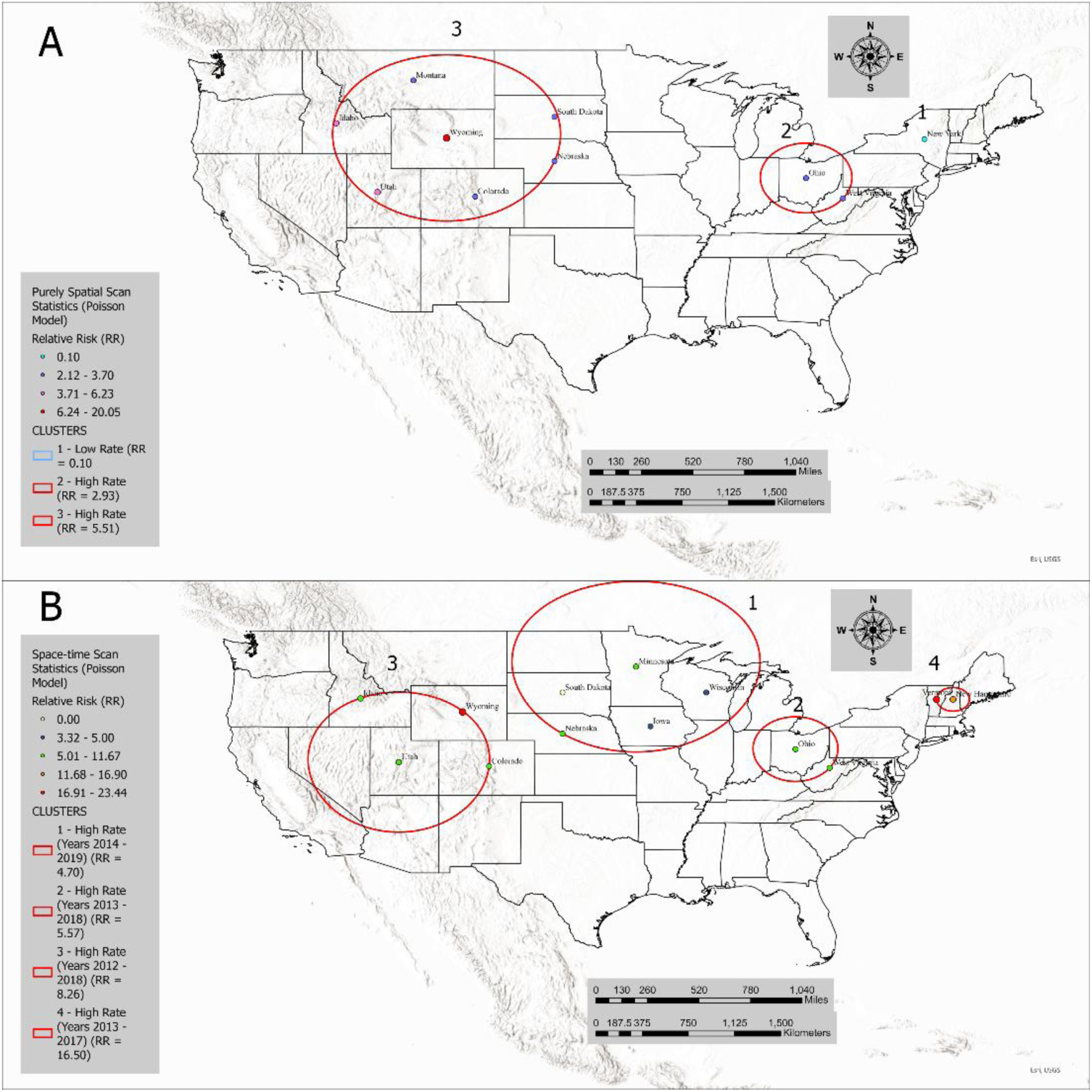
Spatial and space-time clustering of outbreak incidence rates across the U.S. from 2009 to 2022. (A) Purely spatial scan statistics results. (B) Space-time scan statistics results.

The primary spatial cluster was a low-rate cluster that included the state of New York with an RR of 0.10 (Figure 5).

The animal sources that were associated with these outbreak clusters were reptiles (lizards – bearded dragon). The exposure settings were “Other” (Supplemental Table S2).

The second spatial cluster was a high-rate cluster that included two states: Ohio and West Virginia (RR=2.93). The animal sources that were associated with these outbreak clusters were birds (baby chicks and ducklings), reptiles (small turtle, snake, and other reptiles), pet fish, and mammals (cattle, mice, and hedgehogs). The exposure settings were “Other”, school/college/university, and farm/dairy/agricultural settings (Supplement Table S2).

The third cluster was a high-rate spatial cluster that included Wyoming, Montana, Idaho, Utah, Colorado, South Dakota, and Nebraska with an RR of 5.51. The animal sources that were associated with these outbreak clusters were birds (baby chicks and ducklings), reptiles (small turtles, lizards, and snakes), and mammals (cattle, sheep, pigs, goat, dogs, and small mammalian household pets (mice, ferret, hedgehog, and guinea pig)). The exposure settings were farm settings, residential settings, and others.

In space-time scan analysis, four significant clusters that were all high rates were detected using a discrete Poisson model. The primary high-rate space-time cluster occurred between 2014 and 2019 and included five states: South Dakota, Nebraska, Minnesota, Iowa, and Wisconsin, with an RR of 4.70. The animal sources that were associated with these outbreak clusters were birds (baby chicks, ducklings, chickens, turkeys, and ducks), reptiles (turtle, roughneck monitor lizard, and bearded dragon), and mammals (cattle, pig, horse, sheep/goat, ferret, hedgehog, mouse, and guinea pig). The exposure settings were educational institutions, veterinary clinics, and “Other” (Supplemental Table S2).

The second high-rate space-time cluster occurred during 2013 – 2018, and included Ohio and West Virginia with an RR of 5.57. The animal sources that were associated with these outbreak clusters were birds (baby chicks and ducklings), mammals (cattle, pigs, dogs, guinea pigs, and hedgehogs), and reptiles (turtles and lizards). The exposure settings were farm settings, child daycare, educational institutions, and “Other” (Supplemental Table S2).

The third high-rate spatial cluster was located in Idaho, Colorado, Utah, and Wyoming between 2012 and 2018, with an RR of 8.26. The animal sources that were associated with these outbreak clusters were birds (baby chicks and ducklings), reptiles (lizards (bearded dragon), corn snake, and small turtles), and mammals (dogs, pigs, cattle, sheep/goat, and horse). The exposure settings were farm settings, child daycare/preschool, and a veterinary clinic.

The fourth space-time high-rate cluster was detected in Vermont and New Hampshire from 2013 to 2017 with an RR of 16.50. The animal sources that were associated with these outbreak clusters were birds (baby chicks and ducklings), mammals (dogs), and reptiles (pet turtles). The exposure settings were farm settings and “Other” (Supplemental Table S2).

## 4. Discussion

Our study used a stepwise spatial analytical framework, incorporating disease mapping and assessments of global and local clustering of NTS outbreaks linked to animal contact in single states that were reported to the CDC through the National Outbreak Reporting System (NORS) from 2009 to 2022. We examined trends in NTS outbreak incidence rates (IRs), exposure sources, and settings at the state level across two distinct time periods (2009-2015 and 2016-2022). By conducting spatial and space-time scan statistics over the entire study period (2009-2022) to identify higher-than-expected NTS outbreak rates, we assessed the persistence and recurrence of outbreak clusters. This comprehensive approach allowed us to identify states with consistently high-risk clusters and delineate the associated animal contact-related exposure and transmission pathways. The Mountain region (e.g., Wyoming, Idaho, and Colorado) emerged as a consistent high-risk area for NTS outbreaks. This repeated identification across different analytical approaches underscores the robustness of this finding and its relevance to public health.

Our analysis identified differences in animal type and exposure settings of NTS outbreaks across regions. In the Mountain West region, recurrent NTS outbreaks were linked to contact with cattle, poultry, and reptiles on farms, educational, and residential settings, while the Midwest and Appalachia showed diverse host risk factors, including contact with cattle, poultry, reptiles, and small pets. In the Northeast, outbreaks were associated with reptiles and backyard poultry. By contrast, the South region showed persistently low outbreak clustering. Together, these patterns suggest that NTS transmission is shaped by regional animal contact sources, exposure settings, and clustering dynamics, highlighting both persistent hotspots and distinct pathways of zoonotic risk.

In our spatial analysis, global clustering was analyzed across two study periods (2009-2015 and 2016-2022). The first period showed no significant global clustering, while the second revealed a peak at 671.38 km, the distance at which clustering was strongest. Considering the average size of states, this distance matches distances between two adjacent states’ centroids, indicating a clustering at a relatively small extent involving neighboring states rather than broader regions. The NTS burden was more pronounced in the second period (2016-2022), with elevated IRs and recurrent high-high clusters, especially in the Mountain West region.

The spatial analysis consistently identified the Mountain West region, specifically the states of Wyoming, Montana, and Colorado, regions with high livestock density (USDA, 2024), as a high-risk NTS outbreak area. These outbreaks were commonly linked to contact with cattle. Infected cattle, whether symptomatic or asymptomatic, can harbor NTS in their gastrointestinal tract, contaminating their feces and the surrounding environment, which can lead to human exposure through handling livestock or contact with contaminated surfaces, equipment, or water sources (Bentum et al., 2025). Cattle as reservoirs for NTS present challenges for public health, especially in areas with concentrated livestock farming (Arnold et al., 2023). Rural areas with high-density livestock farms have been described previously as risk factors for the transmission of NTS to humans (Shaw et al., 2016). Also, NTS outbreaks and human exposures often originate on farms with improper biosecurity measures (Kudirkiene et al., 2020) where improper handling of animal waste, inadequate sanitation, and poor management of feed and water sources exist (Koyun et al., 2023).

Mitigating the risk of salmonellosis on cattle farms requires developing prevention and control programs, including improved farm management practices, regular health screening of livestock for infections, and educational efforts for both farm workers and the general population to reduce exposure risks (Barnhardt et al, 2025; Shrestha et al., 2025).

The Mountain West region is mainly a rural area, and contact with livestock is not limited only to farm workers. Residents could be exposed to animals at fairgrounds, petting zoos, or while visiting a farm. Animals in these settings, including cattle, goats, pigs, sheep, and poultry, can harbor enteric pathogens such as NTS, and they can shed them in their feces and can also contaminate their environment, leading to human infections through direct contact with infected animals or indirect contact by ingestion of contaminated materials (Conrad et al., 2017; Dunn et al., 2015).

The space-time scan statistics, besides the Mountain West region, identified regions in the Midwest (Ohio, Wisconsin, Iowa, Minnesota, Nebraska) and the Northeast (Vermont and New Hampshire) as high-risk regions over a relatively long period (2013-2019), suggesting the outbreaks are not isolated incidents but part of sustained risk factors in these areas. In our analysis, these NTS outbreaks were associated with contact with livestock (cattle, horses, and pigs) and poultry (baby chicks, ducklings, chickens, and ducks), and the cases’ exposure settings included veterinary clinics, fairgrounds, and farms. This finding could be explained by this region high livestock farm density (USDA, 2024), which increases residents’ exposure risk to infected animals or to their contaminated environment, resulting in recurring outbreaks or disease occurrences over a long period. These environments could facilitate close human-animal contact, and residents’ lack of awareness of zoonotic transmission potential increases their NTS infection risks (Silva et al., 2014). Also, the Midwest is a central hub for livestock production and processing in the U.S, and the movement of animals across state lines can facilitate the spread of pathogens (Cabezas et al., 2021).

Besides, livestock and poultry, contact with reptiles was the third most common NTS outbreak source. Reptiles, including turtles, lizards, and snakes, are recognized reservoirs for NTS, and they can be transmitted to humans through direct or indirect contact. A recent meta-analysis estimated the global prevalence of *Salmonella* spp. in reptiles at 30.4%, with snakes at 63.1% having the highest prevalence, followed by lizards (33.6%) and turtles (11.2%) (Muslin et al., 2025).

Before interpreting our study results few limitations should be noted. Our data included NTS outbreaks that were reported voluntarily by state, local, and territorial health departments, and reporting could be influenced by local public health resources (Tilashalski et al., 2022). Furthermore, we only included single-state NTS outbreaks to assess local exposure sources and settings; however, multistate NTS outbreaks also pose a high public health burden (Eisenstein et al., 2025), and future studies should assess their impact. Lastly, the COVID-19 pandemic was likely a confounder for the decreased NTS reports between 2019 and 2021, as local public health resources were reallocated towards addressing the pandemic (Zarella et al., 2024).

## 5. Conclusion

Our study identified persistent spatiotemporal clustering of NTS outbreaks across the US, consistently identifying high-risk areas in the Mountain West, Midwest, and Northeast of the US. Exposure to cattle, poultry, and reptiles was the most common source of these outbreaks. A multitude of exposure settings, including farms, veterinary clinics, fairgrounds, residential homes, and education facilities, were reported, suggesting that the risk of salmonellosis is elevated in a variety of settings where human-animal interactions are frequent. Locally focused prevention and control strategies are needed in high-risk areas, including the development of public education programs on the zoonotic risks of NTS, restrictions on animal contact, effective sanitation in fairgrounds and petting zoos, and regular surveillance to prevent zoonotic infections.

## Data Availability

All data produced in the present study are available upon reasonable request to the authors.

https://www.cdc.gov/beam/dashboard/index.html

## Acknowledgements

We would like to acknowledge the Centers for Disease Control and Prevention (CDC) and the National Outbreak Reporting System (NORS) for providing access to the dataset. The findings and conclusions in this report are those of the authors and do not necessarily represent the official position of the Centers for Disease Control and Prevention.

## Ethical Approval Statement

The study didn’t require ethical approval.

